# Predicting cause of death from free-text health summaries: development of an interpretable machine learning tool

**DOI:** 10.1101/2021.07.15.21260082

**Authors:** Chris McWilliams, Eleanor I. Walsh, Avon Huxor, Emma L. Turner, Raul Santos-Rodriguez

## Abstract

**Purpose:** Accurately assigning cause of death is vital to understanding health outcomes in the population and improving health care provision. Cancer-specific cause of death is a key outcome in clinical trials, but assignment of cause of death from death certification is prone to misattribution, therefore can have an impact on cancer-specific trial mortality outcome measures.

**Methods:** We developed an interpretable machine learning classifier to predict prostate cancer death from free-text summaries of medical history for prostate cancer patients (CAP). We developed visualisations to highlight the predictive elements of the free-text summaries. These were used by the project analysts to gain an insight of how the predictions were made.

**Results:** Compared to independent human expert assignment, the classifier showed >90% accuracy in predicting prostate cancer death in test subset of the CAP dataset. Informal feedback suggested that these visualisations would require adaptation to be useful to clinical experts when assessing the appropriateness of these ML predictions in a clinical setting. Notably, key features used by the classifier to predict prostate cancer death and emphasised in the visualisations, were considered to be clinically important signs of progressing prostate cancer based on prior knowledge of the dataset.

**Conclusion:** The results suggest that our interpretability approach improve analyst confidence in the tool, and reveal how the approach could be developed to produce a decision-support tool that would be useful to health care reviewers. As such, we have published the code on GitHub to allow others to apply our methodology to their data (https://zenodo.org/badge/latestdoi/294910364).

## 1. Introduction

Free-text electronic health records (hereafter, health records) contain information about a patient’s medical history. In health care, it is common for human experts to review health records to inform decision making and clinical practice. This review process can be time consuming and prone to error and there is considerable potential for algorithmic methods to support human decision-making in this context (1). One example of this process is retrospective auditing of health care practice, which often requires trained experts to manually review and assign outcomes or labels to the cases reviewed. The high cost in person-hours can make such studies prohibitively expensive, including clinical trials, which are essential for improving the delivery of patient care.

As one of the most common cancers diagnosed in the UK, it is not surprising that prostate cancer is one of the cancers on which text mining of health records for research has focused (2). Much of this work has concentrated on predicting cancer detection (3), disease progression (4) and optimizing treatment (5). Knowledge of underlying cause of death is another key health outcome and machine learning techniques have made it possible to extract cancer mortality directly from Medical Certificates of Cause of Death (MCCD)(6). But these methods have not taken into account the inherent misattribution that exists within the death certificate. For example, prostate cancer deaths can be misattributed to other causes which result in underestimates of prostate cancer as a cause of death. This is the case when deaths are attributed to the complications from investigations or treatment of prostate cancer, rather than the disease itself (e.g. infection from biopsy, post-surgery complications). Deaths from other causes can also be attributed to prostate cancer which would cause an over-estimate of prostate cancer as cause of death (7). This was demonstrated in the Cluster randomised trial of PSA testing for prostate cancer (CAP) where an independent committee assigned cause of death, finding that death certification produced false positive prostate cancer deaths 9% of the time. This increased to 23% if the individual had another cancer (not prostate cancer) diagnosed during their lifetime (8). It led to a recommendation that assignment of prostate cancer death, especially as an outcome in trials research, should be confirmed by an independent expert committee (8). In the CAP trial, semi-structured free-text summaries of a patient’s medical history from hospital records are created by trained fieldworkers, which are then reviewed by an independent committee of experts and assigned as either prostate cancer related death or not. The CAP dataset thus provides a binary classification task – the identification of prostate cancer death – for which machine learning algorithms can be trained on the annotation of human experts.

There have been significant advances in the field of text mining in recent years, with general purpose deep neural network models such as BERT (9) achieving state-of-the art performance on multiple natural language processing (NLP) tasks, such as question answering and named entity recognition. Applying such NLP methodologies to clinical text data presents various challenges (10) and perhaps the most significant is the shift in word distributions as compared to the standard corpora on which models are trained. This challenge can be overcome by training on general biomedical corpora (11) and/or by developing task-specific models. For example, the state-of-the-art in the detection of medical concepts, such as ICD-10 codes, is to use task-specific recurrent neural network architectures (12,13) which are pre-trained on a medical corpus such as MIMIC (14). However, on specific tasks, classical NLP approaches are still able to compete with deep learning methods (15–17).

This work addresses the task of document classification. An extensive review of this topic, in relation to clinical text data, is provided by Mujtaba et al (10). Specifically, we aim to train a binary classifier on human expert annotations in order to identify patients that died from prostate cancer using the CAP health records described above. Although optimal performance in such tasks is likely to be achieved by deep learning models, we are more concerned here with the interpretability of the classifier in the context of decision-support. Interpretability is key to the safe and effective deployment of machine learning (ML) in contexts where there is a human-in-the-loop (18). The related concepts of *fairness* (19), *accountability* (20) and *transparency* (21) have become key tenets of ethical artificial intelligence and are nowhere more relevant than in the medical domain. Human users of an ML algorithm must be able to engage with and understand its predictions. Crucially, the user must have the confidence to either accept or reject the predictions based on their clinical expertise and a clear understanding of how the predictions were made.

In this study we developed graphical methods to explain which textual elements contribute to the classification of a health records from CAP by a machine learning algorithm. We also used methods from the FAT Forensics toolbox (22) and the TreeInterpreter Python package (23) to quantify feature contributions. These contributions were then displayed to the user in an interpretable and visually engaging format. The classifiers and interpretability methods were developed for the CAP dataset, using expert committee assignment of prostate cancer-related death to train the classifiers. However, the intention is that these methods will be developed into a decision-support tool that would help human experts in the time-consuming task of classification of health records, both for this specific task and for similar tasks with different data sources.

## 2. Materials and Methods

The machine learning classifier was developed using the CAP dataset, following which we investigated a variety of visualization techniques to aid interpretation of the ML predictions. The CAP dataset is outlined below (2.1). All code was written in Python and has been released publicly at https://zenodo.org/badge/latestdoi/294910364 to facilitate re-use by other groups working in this area.

### 2.1 CAP medical history summaries

The CAP trial is a cluster-randomised control trial (RCT) that aims to investigate the effectiveness of screening for prostate cancer in the UK population, which has been running since 2001. The CAP dataset used in this study contains 2,606 medical summaries of men who had a diagnosis of prostate cancer, registered with the UK National Cancer Registrations Service in England and Wales, and who had died by 31^st^ March 2016 (see Table 1 for characteristics). Deaths were notified by the Office of National Statistics who provide death certification information via NHS Digital.

**Table 1.** Characteristics of prostate cancer patients in the CAP dataset

| Characteristic | Prostate cancer dataset (CAP) |
| --- | --- |
| No. of patients | 2,606 |
| Sex | M: 2,606 (100%); F: N/A |
| Age at diagnosis (mean) | 67.5 years (IQR: 64.1-71.3) |
| Age at death (mean) | 70.2 years (IQR: 66.5-74.5) |
| Year of death (range) | 2003 - 2016 |
| Primary outcome | Prostate cancer related death<br>Yes: 1,095 (42%); No: 1,511 (58%) |
| Note types | Oncology and urology notes, inpatient notes, outpatient notes, radiology notes, multidisciplinary team meeting notes, lab reports, operative reports |

The medical summaries were written by 13 trained fieldworkers who extracted prostate cancer and end of life information directly from the medical records in hospitals. The semi-structured summaries of the patients’ medical history include information about prostate cancer severity, treatments, and progression, as well as co-morbidities and competing causes of death. The full list of text fields present in this data are given in table ST1 in Online Resource 1.

The medical summaries have been reviewed by an independent cause of death evaluation committee made up of clinical experts (i.e. urological surgeons, pathologists and oncologists) who decide whether the cause of death is due to prostate cancer for each case, or whether the patient died ‘with’ prostate cancer and not ‘from’ prostate cancer. This human made decision was used as the ground truth against which to train the classifier.

### 2.2 Machine learning pipeline

We used a bag-of-words representation of the health records which was produced as follows. The semi-structured text fields were concatenated into a single string and then lemmatized using the *WordNetLemmatizer* from Python’s Natural Language Toolkit (NLTK). The full details of the lemmatization are provided in Algorithm A1 in Online Resource 1. We then extracted features from the lemmatized text using *CountVectorizer* and *TfidfTransformer* from Scikit-learn. The hyperparameters of *CountVectorizer* were optimized using *GridSearchCV* along with those of the classifier algorithm. The classifier was trained on 80% of the dataset with 20% held out for testing. Following optimisation of the hyperparameters with 5-fold cross validation, the best classifier was refitted to the full training data. Full details of the feature extraction and model training are provided in Algorithm A2 in Online Resource 1. During model development we tested three classifier algorithms from scikit-learn. However, the code can easily be adapted to use any off-the-shelf or bespoke classifier. We evaluated classifier performance using a suite of metrics, including accuracy and AUC (area under the curve).

### 2.3 Interpretability methods

On top of the classification pipeline, we developed interpretable outputs that allow users to engage with the classifier’s prediction and which are intended to act as a proof-of-concept for a future decision support tool based on this work. To achieve interpretability, we focused on the communication of *feature importance*. We intended to convey an intuition for how the classifier works and to allow users to consider whether they agree with individual predictions based on how that prediction was made. In other words, we introduced the key elements of *accountability* (20) and *trust* (18) to the classification system. We used word clouds to visually represent the relative importance of the features (words and bigrams) by scaling their size. The colours of the words were used to display the sign of the contribution of the feature towards the class prediction (where this information was available). We extended this visual representation by producing augmented versions of the original free-text summaries that displayed feature contributions in the context of the original language used.

To quantify feature importance, we used four different approaches. Two of these methods are specific to the tree-based classifiers, while two are generally applicable and can be used with any machine learning classifier. The four methods are as follows:

1. *Gini Importance* – Is calculated as the normalised total reduction in the Gini impurity brought about by splits on a given feature across the ensemble of trees (24). This metric is part of the scikit-learn implementation of the Random Forest classifier and only provides an aggregate measure of importance (i.e. at the level of the whole dataset). It also does not capture the sign of the contribution of the feature.
2. *TreeInterpreter* – This package (23) computes feature importance by decomposing a single prediction into a sum over feature contributions. It captures both the sign and magnitude of each feature contribution to a single prediction and can be averaged over many predictions to produce an aggregate measure of importance.
3. *LIME –* Local Interpretable Model-agnostic Explanations (25) are a standard tool of explainable AI and are implemented in (22). They provide a local measure of feature contribution that can be used with any machine learning classifier.
4. *SHAP -* SHapley Additive exPlanations (26) is a model agnostic explainer based on game theoretic methods that also provides a local measure of feature importance.

## 3. Results

A random forest classifier proved to be the best performing algorithm for the task of predicting prostate cancer death according to accuracy and F1 score. This model achieved a classification accuracy of 91.8% and an F1 of 0.931 on the test data. (Its cross-validation performance is illustrated in figure S0 in Online Resource 1.) A support vector classifier and linear regression classifier achieved comparable levels of performance (see Table 2 and Figure 1). We attempted recalibration of the random forest classifier output using both isotonic and sigmoid (27) methods (see Figure S1 in Online Resource 1) but both methods made only minor improvements to the probabilities output. As such, the main results presented here are for the uncalibrated classifiers. We hypothesized that the cases represented in the CAP dataset exhibited different degrees of classification difficulty, corresponding to the method with which the cases had been originally labelled with a cause of death. These different methods are collectively referred to as the “*cause of death route”* (COD) and are explained in Algorithm A3 in Online Resource 1. Using the COD route, we stratified the dataset into *hard* and *easy* cases. We found that the classifier performance was worse for the hard cases than the easy cases (Figure S2 in Online Resource 1) and that the probabilities were better calibrated for the easy cases (Figure S3 in Online Resource 1). For hard cases, the classifiers tended to underestimate the probability of prostate cancer death towards the lower end of the range. This implies the existence of patients who actually died of prostate cancer, but which look to the classifier like they did not. These results suggest that the stratification of cases based on COD route is meaningful and may have significant implication for how a decision support tool could be used and evaluated in the future. However, it should be noted that these groups are imbalanced with 2340 and 270 easy and hard cases appearing in the dataset, respectively. We investigated the effect of authorship and found evidence of clustering based on language style (Figure S4 in Online Resource 1) but this did not significantly affect performance (see section SM1.1 in Online Resource 1).

**Table 2.**
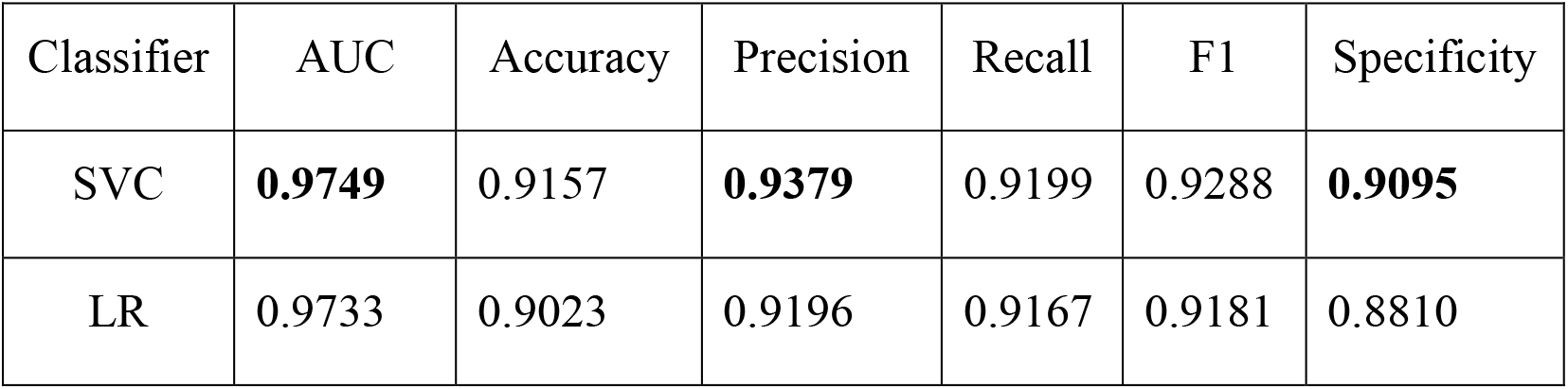
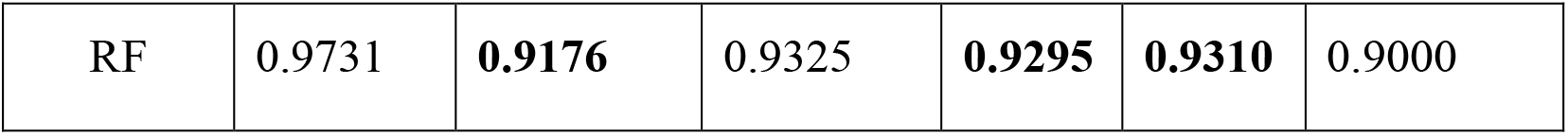
Performance summary of the three classifiers. SVC = Support Vector Classifier; LR = Logistic Regression; RF = Random Forest. Bold face indicates the best performance for each metric.

**Figure 1:**
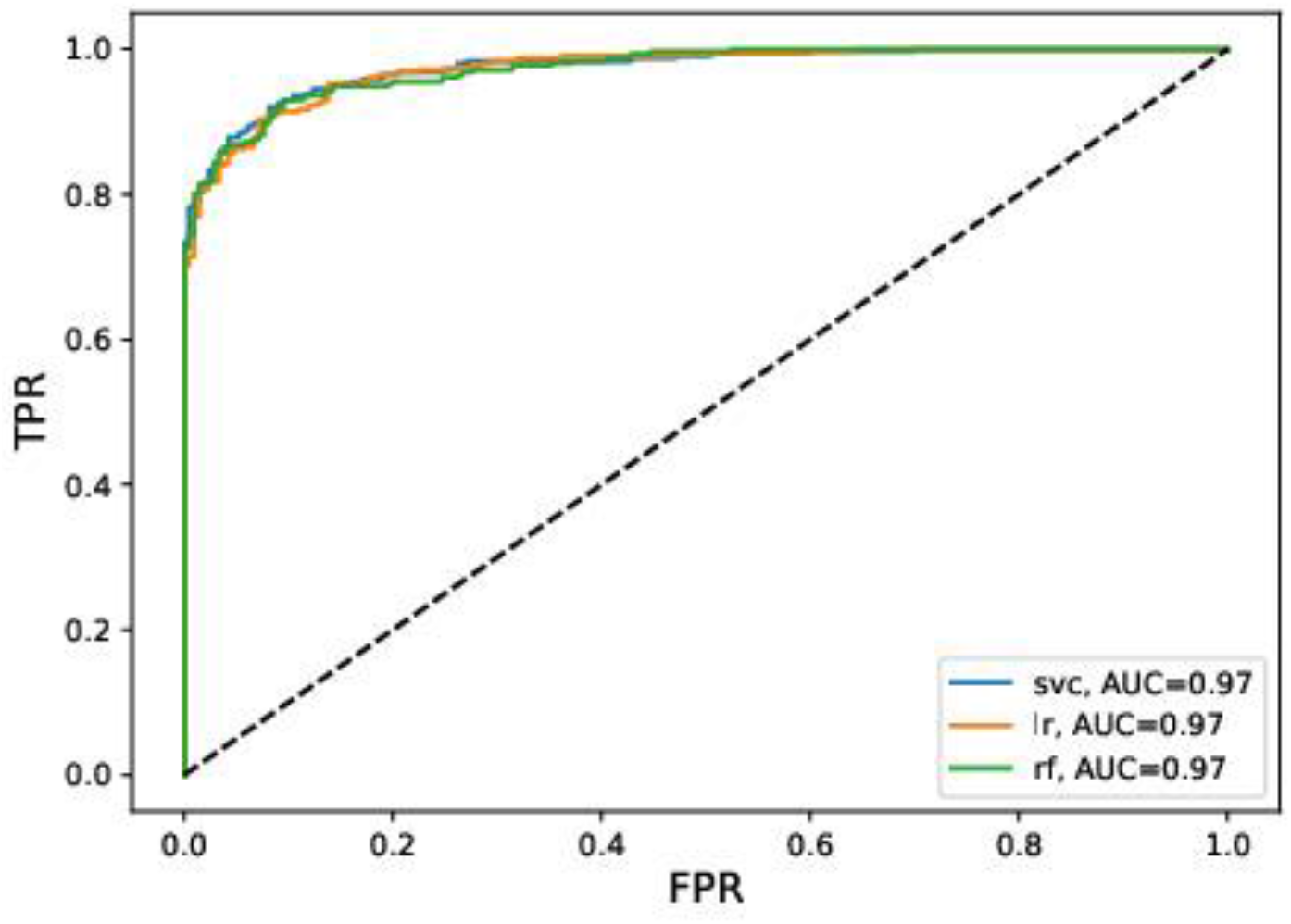
Receiver operating characteristic (ROC) curve for three classifiers predicting cause of death from the CAP dataset, plotting the true positive rate (TPR) against the false positive rate (FPR). Performance curves are shown for: SVC = Support vector classifier; LR = logistic regression; RF = random forest

The random forest was chosen to explore interpretability based on its performance. This choice also allowed us to explore feature importance metrics that are only applicable to tree-based algorithms. We compared feature rankings obtained using the four metrics for feature importance using Spearman’s rank correlation coefficient (see table ST2 in Online Resource 1). The rankings according to SHAP, LIME and Gini importance were all moderately to strongly correlated with the TreeInterpreter rankings (ρ = 0.54 - 0.65). Here we present interpretability results using the TreeInterpreter metric for feature importance, but the equivalent outputs can easily be produced using the alternative metrics (see for example, Figures S5 and S6 in Online Resource 1).

We produced word clouds to illustrate the most important features that contributed to the classifier predictions. These word clouds were shared with members of the CAP study team, who confirmed that the classifier was using clinically meaningful information to make predictions. Features with large contributions (such as “bone scan”, “spine”, “hormone”, “androgen”) tend to be associated with advanced stage prostate cancer. These features can contribute positively or negatively to the classification depending on the frequency of occurrence of the term in the health record. For example, both “hormone” and “bone scan” contribute positively to the classification of prostate cancer death when present in individual cases (see Figure 2(B)) but when averaged across the dataset they are indicative of non-prostate cancer death (see Figure 2(A)).

**Figure 2:**
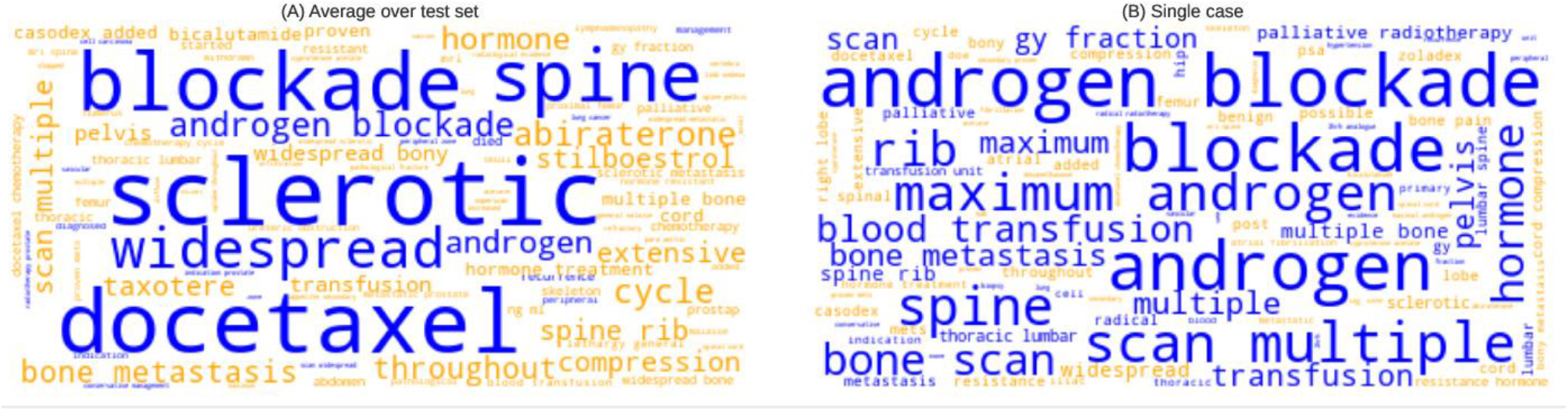
Word clouds indicating the feature contributions towards random forest predictions of prostate cancer death (for the classifier depicted in Figure 1). The size of the word or bigram indicates its relative importance. Blue words or bigrams are indicative of prostate cancer death, while orange is indicative of not prostate cancer death. Feature contributions determined using TreeInterpreter (see main text). (A) Average feature contributions over the CAP test set; (B) a single case that was correctly predicted prostate cancer death by the classifier (shown in Figure 3)

Therefore, we felt it was necessary to see these feature contributions in the context of the original text, to determine if the classifier is correctly identifying textual elements that indicate prostate cancer death. For this reason, we sought a format that would allow users to engage with the classifier output and could potentially be used for decision support. The solution, arrived through dialogue with members of the CAP project, was to produce augmented versions of the original health records which we refer to as *interpretable vignettes*. A partial view of one of the interpretable vignettes is shown in figure 3 and full examples are provided in Online Resources 2 and 3. The text uses the same formatting as the word clouds to show feature contributions. Here the reader can see the context in which the features appear and can therefore use their judgement to determine if the classifier is using the textual element in a meaningful way towards its prediction (28). A legend is provided so that readers can interpret the relative feature contribution sizes and summary information is provided in the header of the vignette.

**Figure 3:**
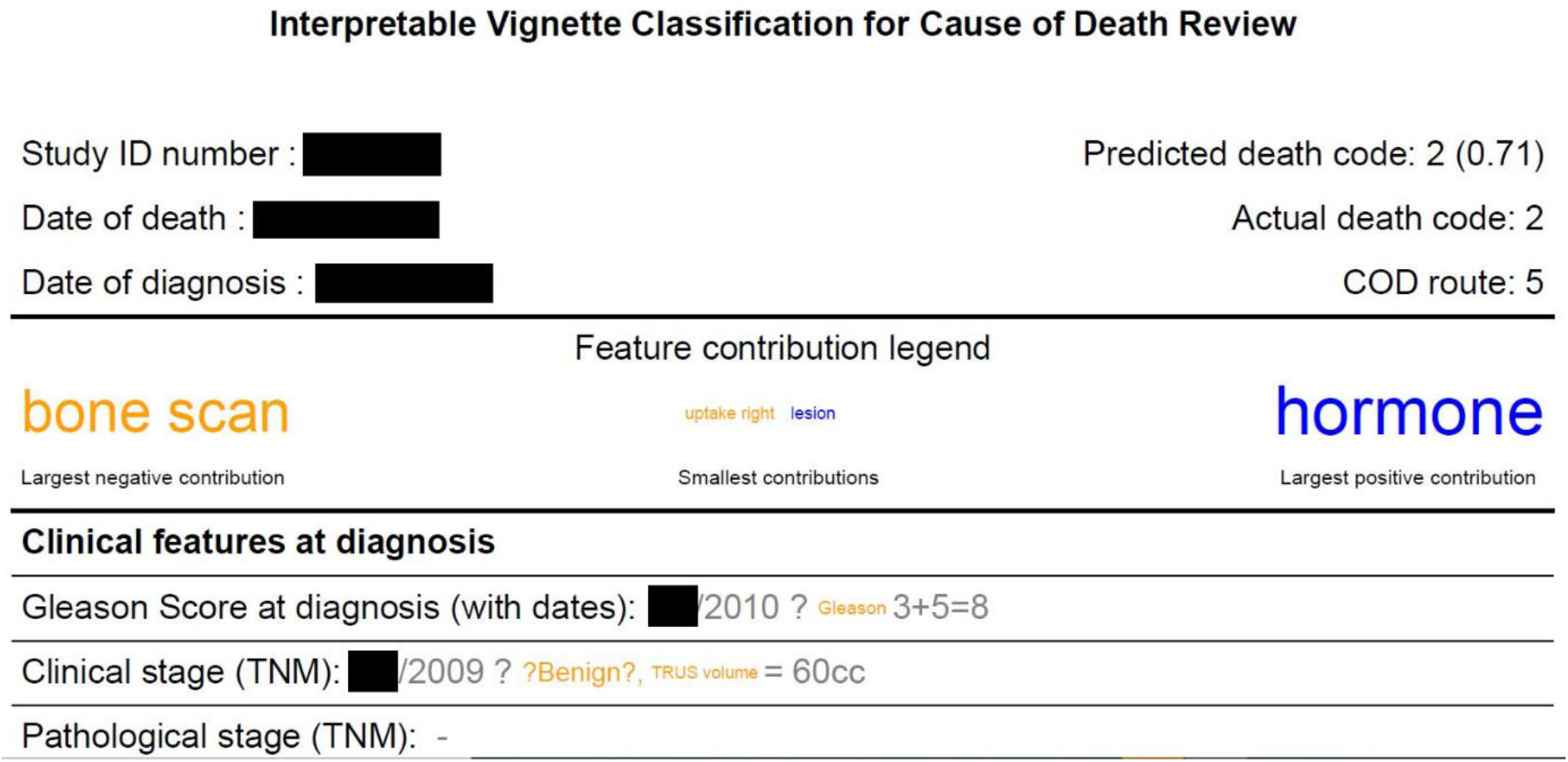
Snapshot of an ‘interpretable vignette’ that allows users to engage with the prediction that is made by the classifier. This case was correctly predicted to be a prostate cancer death by the classifier (cause of death code = 2). As in figure 2 the word (or bigram) size indicates the magnitude of the contribution of that feature to the prediction and the colour indicates the sign of the contribution. Here the original format of the vignettes is retained, which is the format in which the decision makers would normally engage with the document. Full interpretable vignette examples are provided in Online Resources 2 and 3

## 4. Discussion

For a machine learning tool to be useful, especially in the medical domain, it is essential for the user to be able to interpret its output. As such, it is common for studies about clinical decision support or prediction modelling to include a discussion of interpretability (29–31). However, as Lipton points out, “interpretability is not a monolithic concept” (32) – it includes distinct yet intersecting ideas such as comprehension, transparency and trust. It is also subjective, in that what one user may find interpretable another user may not. Most importantly, what the data scientist may consider to be interpretable is not necessarily of use to a clinical decision maker. In this study we used four standard interpretability metrics to produce measures of feature importance from a machine learning classifier that was trained to identify prostate cancer deaths from medical summaries. We then developed visual representations of these feature importances and presented them to one of the trained CAP reviewers, who would be the intended users of a future decision support system based on this work. Their feedback is summarised below and used to highlight the strengths and weaknesses of our approach, and to identify directions for future work.

The word clouds of feature contributions were intended to provide a high-level overview of what the classifier learned from the data. The reviewer showed a preference for an alternative presentation of this information, stating that “they may be a reasonable way to display the model weights, but I probably would better understand a sorted list in tabular form with associated weight magnitudes”. The augmented vignettes (Online Resources 2 and 3) proved to be more useful. These allowed the reviewer to engage with individual predictions, by highlighting features in the original text, using font colour and size to indicate the direction and magnitude of the contribution to the prediction. Similar visualisations have been used in other ML studies to provide interpretability (28,33–35). Being able to see the feature contributions in the semantic context of the original text enabled the reviewer to determine where the classifier was correctly or incorrectly using features. In general, the reviewer felt that the highlighted feature contributions were consistent with their clinical assessment:

> *“Many of the colors make sense here (in Online Resource 2). Metastases would be consistent with prostate cancer death…Meanwhile, mentions of “lungs”, “ascites”, “stomach”, and “thorax” in the vignette suggest the patient has some non-prostate-cancer condition that is worthy of attention—those are appropriately yellow*.*”*

The features appearing in Online Resources 2 and 3 which were associated with prostate cancer death, aligned with the indicators of advancing disease (e.g. bony metastases, hormone treatment) that are used as clinical outcomes in prostate cancer trials (36). However, the feedback made it clear that the visualisations were less interpretable than intended:

> *“I am confused, again, that ‘bone scan’ and ‘hormone’ are blue here (in Online Resource 3) but were each yellow elsewhere (bone scan was yellow for Online Resource 2 and in Figure 2A; hormone was yellow in Figure 2A)*.*”*

Here the reviewer is referring to the ability of a feature (e.g. ‘bone scan’) to contribute positively to a classification of prostate cancer death when present in the text, but to contribute negatively to the classification when it is absent. This is an example of the potential for conflict, referred to by Lipton (32), between what is a transparent and faithful representation of the mechanism of a classifier and what is easily understandable by a human user. In this case, the problem might be overcome either by including some indication of the actual feature value or by providing some training to the user to resolve the apparent inconsistency.

The augmented vignettes let the user see if elements of text are missed or used incorrectly by the classifier, in which case they can exercise caution when considering the prediction. In this way the reviewer determined:

> *“that some of the descriptions I pay most attention to (the rising PSA values and, to a lesser extent, the high Gleason score) are gray—presumably because the algorithm is ignoring them*.*”*

Both PSA and Gleason score are numerical values which are often included in these medical summaries, but which are not captured by our current feature representation. This feedback suggests that an avenue for improved performance would be to incorporate prior clinical knowledge such as the importance of these two scores. Interestingly, it may also improve trust in the system if users could see that the classifier was making use of the elements of the medical summaries that they consider to be most important. The interpretable vignettes also revealed that classification of prostate cancer death was problematic when *negation* appeared in the text. Our bag-of-words feature representation would not be expected to handle negation, so the application of methods to detect negation in clinical text data (37,38) would likely boost performance. Off-the-shelf classifiers achieved good performance on the CAP dataset. For different health record datasets, additional effort may be required to achieve sufficient performance for a decision support tool to be useful. The clustering of the health records based on authorship suggested that methods such as multiple-source cross-validation (39) or domain adaptation (40) could be beneficial in dealing with differences in writing styles within other datasets. Other methods to boost performance would likely be task- or domain-specific and could include the addition of numeric clinical features extracted from structured data (41), or the use of state-of-the-art deep learning methods (12,37). Such methods would be compatible with our model-agnostic approach to interpretability.

The CAP dataset contains a proxy for ‘difficulty’ of the cause of death assignment. Although we were not able to train a model to reliability predict hard cases, our cause-of-death-classifier did show worse performance and calibration on the hard cases than on the easy cases. This suggests that the stratification of cases according to difficulty is meaningful and is likely to have implications for the future development and evaluation of a decision support tool. A systematic investigation of what makes the hard cases more difficult to classify, and which features are most predictive for different types of cases, will help to inform more targeted data acquisition from hospital records. Named-entity recognition approaches could also be adapted to assist with this information retrieval (12,15). Such knowledge could produce significant cost savings in data collection for CAP and similar projects. In practice, the predictions for hard cases are less trustworthy and one way to address this would be to produce reliable estimates of uncertainty (42). In an applied setting it would be important for the transparency of the system to communicate to users the relative risks of both false positive and false negatives.

The feedback of the CAP reviewer has given us confidence in the feasibility of these methods, and the next stage is to develop them into a usable decision support tool, following a user-centric design process with members of the intended user group (43). Key to this will be to adapt the visualisations to be appropriate for users in a clinical setting. We will need to test our classifiers on new CAP reviews to determine how well they generalise to unseen data. Our bag-of-words approach is limited by the size of the training data. There are 1360 words in the test data set that do not appear in our training data (Figure S7 in Online Resource 1), and the CAP dataset has only limited overlap (Figure S8 in Online Resource 1) with an example biomedical corpus (44). To optimise classifier performance in the future will likely require an adapted pre-trained deep learning model (11). We have identified benchmarking datasets (45) that would allow comparison of different classification approaches to ensure that the best model can be selected. It is clear from our results that the different approaches to quantifying feature importance produce distinct feature rankings. Choosing the best approaches to ensure user trust and system transparency will be achieved using A/B testing across a range of users. The continued use of model-agnostic explainability methods will allow abstraction of the decision support interface from the underlying classifier and would allow the tool usable across a range of different tasks and datasets. For example, we plan to test our approach to interpretable document classification on an intensive care dataset (46) that contains free-text medical notes that are routinely used by hospital staff to audit clinical practice.

## 5. Conclusion

Algorithmic classification of health records, such as the identification of prostate cancer death in the CAP dataset, could reduce the need for complex medical summaries to be reviewed by an independent committee. We have demonstrated use of visual methods to explain classifier predictions to human users, which could be deployed in a future decision support tool to reduce the cognitive burden on individual reviewers. Knowledge of the predictive features could also be used to target data extraction from hospitals, reducing the workload and cost required in creating the free-text summaries. We encourage researchers to take a user-centric approach when developing interpretable machine learning tools, to ensure maximum trust and usability in the system.

## Data Availability

The dataset analysed in this studied cannot be shared publicly for reasons of data protection.

## Declarations

### Availability of data

The dataset analysed in this studied cannot be shared publicly for reasons of data protection.

### Code availability

Code is available at https://zenodo.org/badge/latestdoi/294910364

### Author Contributions

CM and AH conducted data analysis and modelling. EW, AH, ET and RS developed the study concept and design. EW and ET contributed domain expertise and detailed understanding of the dataset. RS contributed expertise in interpretable machine learning. EW and CM wrote the manuscript and all authors contributed edits and revisions.

## Acknowledgments

We acknowledge the contribution of the CAP trial group, including the Principle Investigators Professor Richard Martin (Lead), Professor Jenny Donovan, David Neal and Freddie Hamdy. A special thanks to the Cause of Death Evaluation Committee for reviewing the summaries written by the fieldworkers Naomi Williams, Siaw Yein Ng, Laura Hughes, Elizabeth Hill, Charlotte Davies, Liz Salter, Jainnee Mauree, Mari-Anne Rowlands, Leah Bowen, Sean Harrison, Pete Holding and Lindsey Bell. And thank you to Tyler Seibert, Assistant Professor and Radiologist at University California San Diego for his insights into the data visualisation outputs from this project. The CAP trial recognises that this work uses data provided by patients and collected by the NHS as part of their care and support. We also acknowledge the work of the NHS Digital Organization and Office of National Statistics for their assistance with this study.

## Ethics statement

This work was completed within the ethical approval already held CAP project (Derby National Research Ethics Service Committee East Midlands & NHS Health Research Authority Confidential Advisory Group (CAG) approvals). In the case of the CAP trial, the fieldworkers are highly trained to include minimal identifiable information (e.g. they exclude person names, hospital identifiers, full dates etc.). A derived extract of the, already pseudonymised, CAP summaries were then analysed.

## Consent to participate

Not applicable.

## Consent to publish

Not applicable.

