## Supplementary material for "Predicting cause of death from free-text health summaries: development of an interpretable machine learning tool"

**Journal of Healthcare Informatics Research**

**Chris McWilliams[1¥], Eleanor I. Walsh[2¥*], Avon Huxor[3], Emma L. Turner[2], Raul Santos-Rodriguez[1]**

[1]School of Computer Science, Electrical and Electronic Engineering, and Engineering Mathematics, Faculty of Engineering, University of Bristol, Bristol, UK

[2]Bristol Medical School – Population Health Sciences, Faculty of Health Sciences, University of Bristol, Bristol, UK

[3]School of Engineering, Mathematics and Physical Sciences, University of Exeter, Exeter, UK

[¥] co-lead authors

*** Correspondence:**
**Corresponding Author**
****

### Section SM1.1

We found a strong clustering based on authorship, as can be seen in figure S4 where each colour represents a different author. We were interested in the 'pink author' which appears to have two distinct styles of language. However, we were not able to identify a cause for this split (hypotheses included a shift in writing style following training or some other temporal variation). It is possible that the author ID was misattributed to some of these health records.

When we trained the classifier on a only subset of the authors and tested it performance on records written by an author that was not in the training set, we did not see a noticeable drop in performance. This suggests that, although there are clear differences in writing style between authors, that classifier is able to handle them.

### Section SM1.2 - Alternative measures of feature importance.

The results presented in the main text used the Python package TreeInterpreter to determine the contributions of each feature for any given prediction. In section 2 we defined two feature measure of feature importance: 1) Gini importance and 2) LIME.

1) The Gini importance is built in to the scikit learn implementation of the random forest classifier. It is an aggregate measure and therefore only tells us about the classifier (ensemble) behaviour at the level of the whole dataset. It also does not tell us the sign (positive/negative) of each feature contributions. The aggregate feature importances, calculated using this metric, are shown in figure S5.

2) LIME is a good alternative to TreeInterpreter because it can tell us about the feature contributions for a specific prediction by fitting a surrogate model in that (local) region of the feature space. LIME

is model agnostic and therefore could be used with any underlying classifier. Figure S6 shows the same case as Figure 2(B) in the main text, but with feature contributions calculated using LIME.

**Section SM1.3**

As described in the main text, the “cause of death route” was used to stratify cases into *hard* and *easy*. Conceptually the hard cases were those the required more expert input to assign a cause of death based on the health record. We attempted to use the same feature set (that was used for prediction of prostate cancer death) to train a classifier to predict hard cases. The performance of the classifier was poor (AUC~0.6), suggesting that either the features used did not capture what made these deaths hard to assign a cause to, or simply that identifying hard cases is a difficult task in itself.

*Table ST1: List of all the text fields in the CAP dataset.*

| Data field |
|---|
| 'gleason', |
| 'clinical stage', |
| 'path stage', |
| 'comorbidity', |
| 'primaries', |
| 'diagnosis psa', |
| 'local spread', |
| 'diag mets', |
| 'treatment', |
| 'hormones', |
| 'mab', |
| 'orchidectomy', |
| 'chemotherapy', |
| 'complications', |
| 'serial psa', |
| 'testosterone', |
| 'prog mets', |
| 'progression', |
| 'recurrence', |
| 'treat comorb', |
| 'final symptoms', |
| 'final consult', |
| 'ds1500', |
| 'post mortem' |

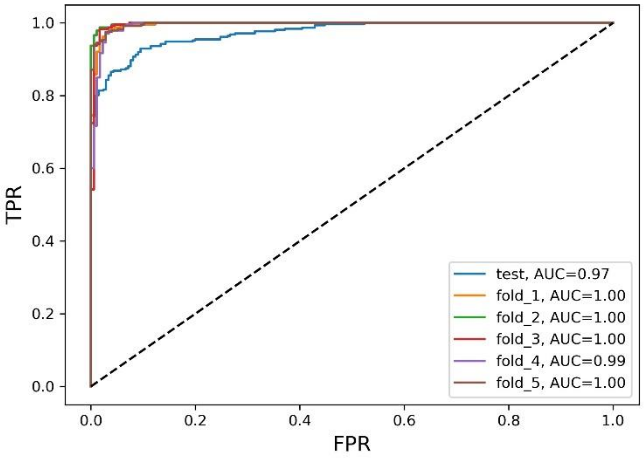


**Figure S0: Receiver operating characteristic curves for the random forest classifier for each of the 5 validation folds used in the cross-validation, compared to the held-out test set. AUC values are given in the legend to 2 decimal places.**

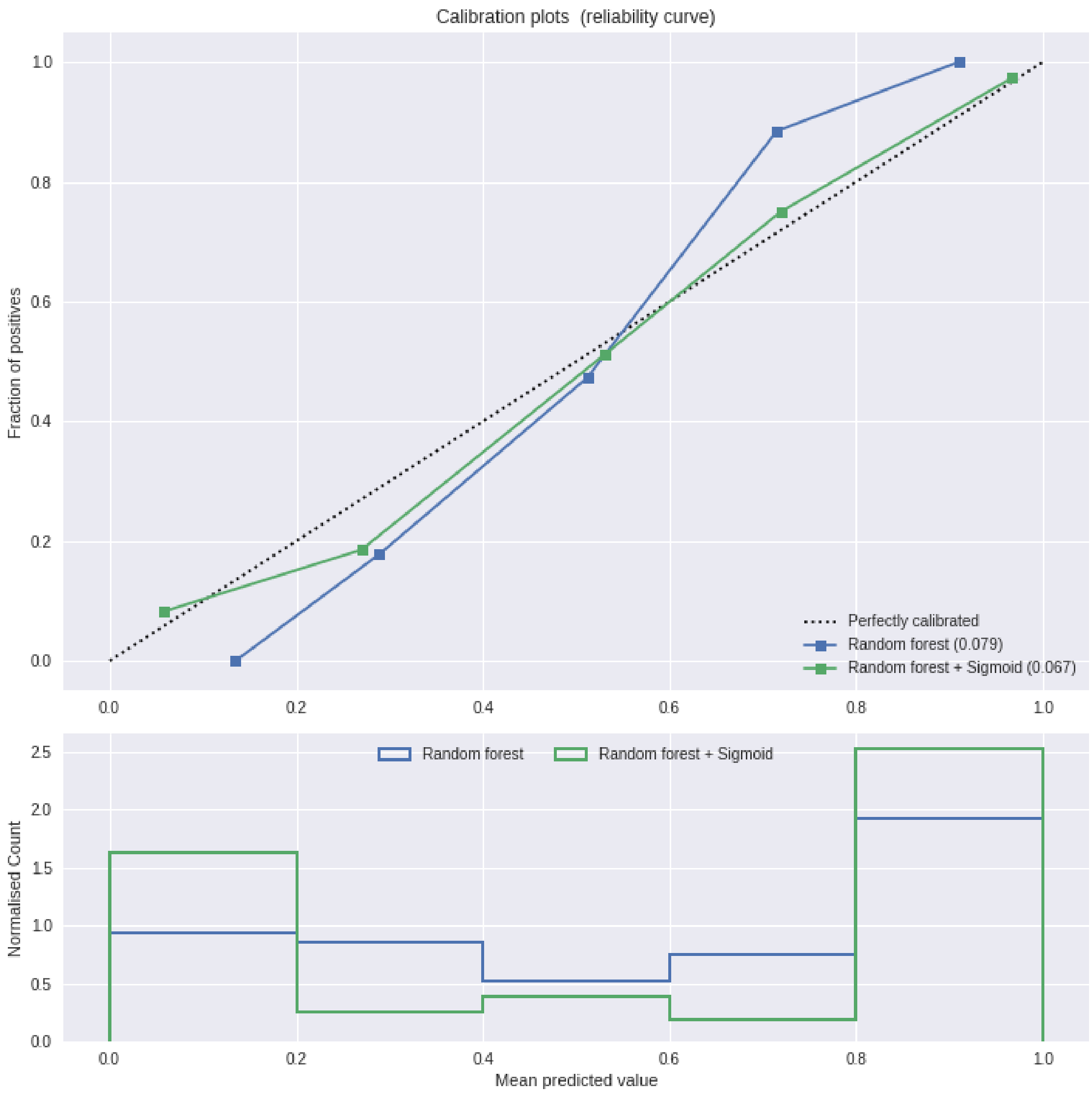


**Figure S1:** ***Calibration curve for the random forest trained to predict prostate cancer deaths in the CAP dataset. To produce these plots, the samples in the test set are grouped into five equally sized bins based on the classifier predictions ('Mean predicted value' is the mean of the classifier predictions within each bin). The 'Normalised count' and the 'fraction of positive' are the number of samples in the bin, and the fraction on samples in the bin belonging to the positive class, respectively. Note that the x-axis is shared. 'Perfectly calibrated' indicates the scenario where the bin means are equal to the fraction of positives in each bin. Points above/below this line indicate***

***that the predictions made by the classifier in that region of probability space are too low/high on average.***

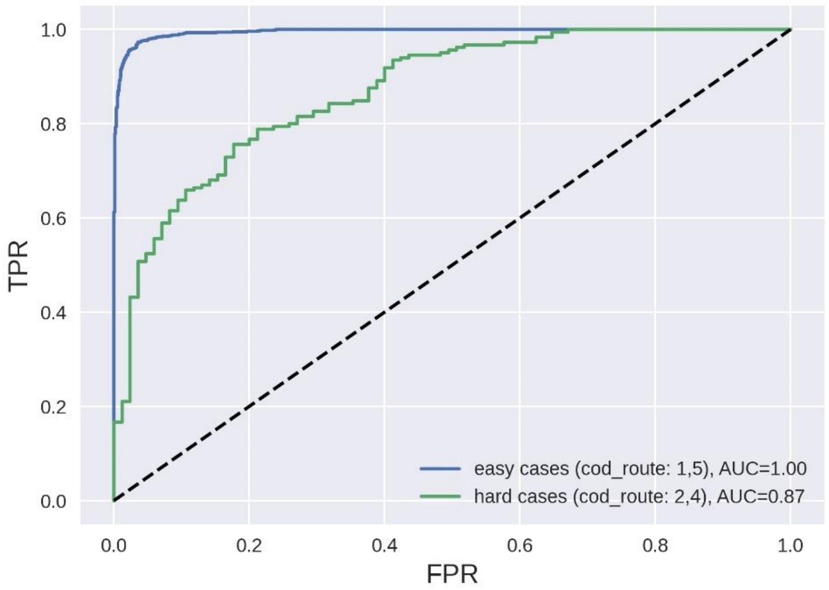


***Figure S2: ROC curve for the same classifier depicted in the left-hand panel of Figure 1 in the main text. Here the performance curves for ‘easy’ and ‘hard’ cases are depicted separately.***

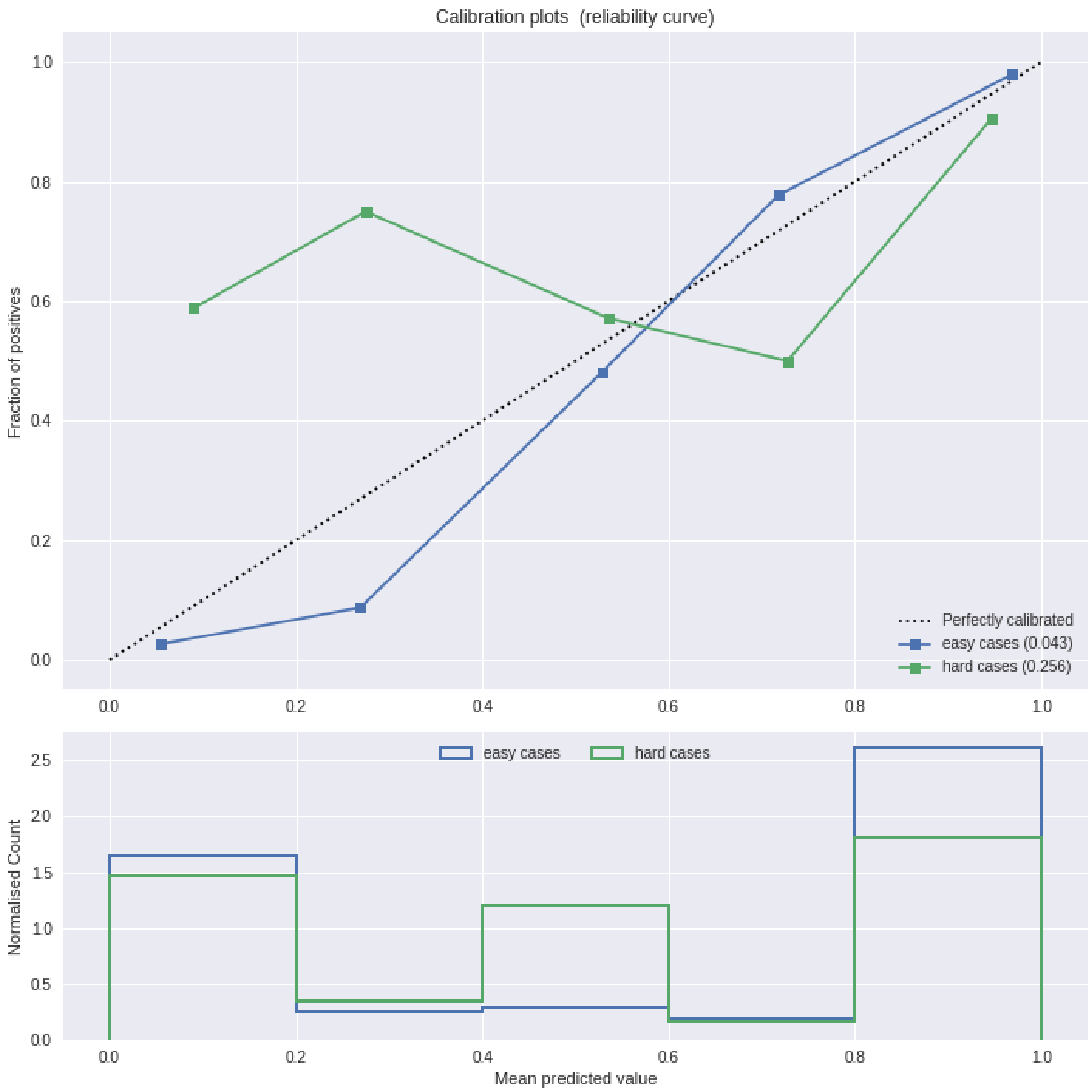


***Figure S3: Calibration curve for the same classifier as figure S1, but with the calibration for hard and easy cases shown separately.***

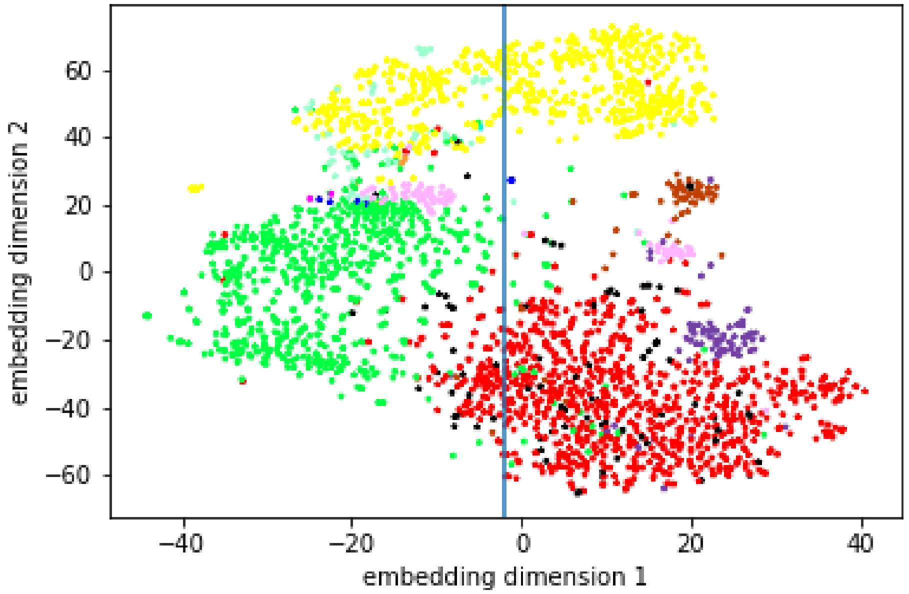


***Figure S4: A two-dimensional t-SNE embedding of the CAP feature space for the classifier depicted in the left-hand panel of Figure 1 in the main text. The cases are coloured by author ID number. There is a clear clustering based on the author of the text. The split in the pink authors cases was investigated (see section SM1.1).***

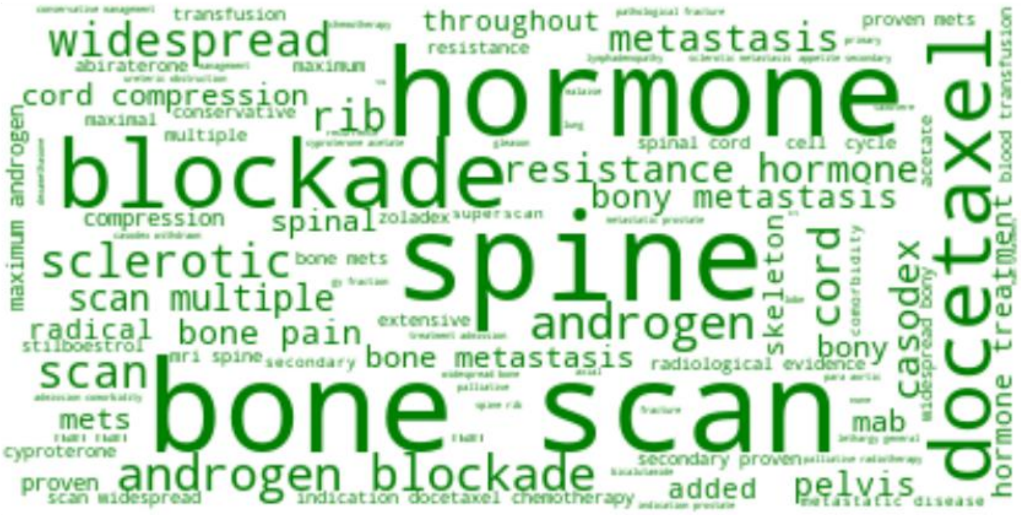


***Figure S5: Word cloud for the same classifier depicted in the left panel of Figure 1 in the main text. Here the feature importances are computed using the Gini Importance (as is standard with the scikit-learn implementation of the Random Forest). This metric provides only the magnitude of importance, not the sign of the contribution.***

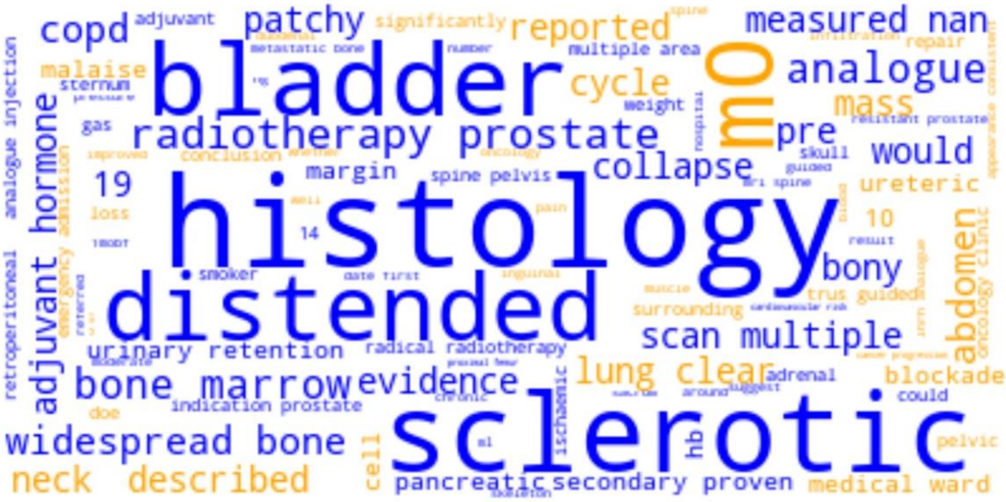


***Figure S6: Word cloud showing LIME feature contributions for a single case (the same as Figure 2B in the main text).***

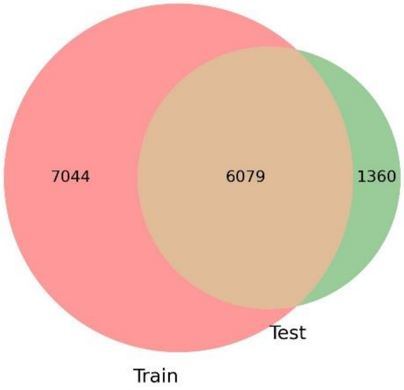


**Figure S7: Number of unique words appearing in the training and test data sets derived from our CAP data.**

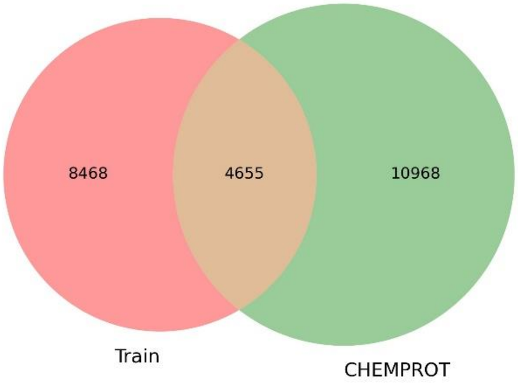


**Figure S8: Number of unique words appearing in our train data compared to the CHEMPROT dataset.**

| feature | rank_ti | rank_shap | rank_lime | rank_rf |
|---|---|---|---|---|
| sclerotic | 0 | 38 | **4** | **6** |
| docetaxel | 1 | **11** | **0** | **5** |
| blockade | 2 | 84 | **2** | **9** |
| spine | 3 | 177 | 20 | **2** |
| widespread | 4 | 20 | **3** | **3** |
| androgen blockade | 5 | **15** | **1** | **8** |
| cycle | 6 | **9** | 40 | 36 |
| abiraterone | 7 | 54 | **12** | 38 |
| androgen | 8 | 13 | **5** | **4** |
| hormone | 9 | **0** | 34 | **0** |
| throughout | 10 | **4** | 38 | 25 |
| bone metastasis | 11 | **12** | 57 | **12** |
| spine rib | 12 | 194 | 72 | 102 |
| extensive | 13 | 21 | 43 | **28** |
| scan multiple | 14 | 91 | **10** | **27** |
|  | **# shared** | **6** | **8** | **11** |
|  | **Spearman R** | **0.599 *** | **0.545 *** | **0.647 *** |

**Table ST2: The top 15 important features according to the TreeInterpreter method, and the corresponding rankings produced by the three other feature importance approaches: SHAP, LIME and RF (the in-built Gini importance of the Scikit-learn Random Forest implementation). The number of the top 15 features shared with between approaches is provided. The Spearman rank correlation coefficient is computer for all 1500 features (* indicates p-value  < 0.01).**

**Algorithm A1: Lemmatization of text**

Input: Dataframe *D* consisting of *c* columns of free-text data, instance *L* of NLTK *WordNetLemmatizer* object.

Uses: Process text data for use in bag-of-words feature representation

Output: List *X* of lemmatized words

Algorithm:

1. Concatenate text from *c* columns into a single string
2. Remove special (non-word) characters
3. Remove single characters
4. Replace multiple spaces with single space
5. Convert text to lower case
6. Loop over words, lemmatize using *L* and append each to list *X*

**Algorithm A2: Feature extraction and model training**

Input: List *X* of lemmatized words (output of A1) and list *y*, instance *C* of Scikit-learn *CountVectorizer* and instance *T* of Scikit-learn *TfidfTransformer*

Uses: Train classification model on clinical text data

Output: Trained classifier model *M* (here a RandomForestClassifier)

Algorithm:

1. Randomly split *X* and *y* into training and test sets (80:20)
2. Create Scikt-learn Pipeline *C* -> *T* -> *M*
3. Define hyper-parameter ranges over which to optimise
4. Run Scikit-learn GridSearchCV with 3-fold cross-validation
5. Return GridSearchCV *best_estimator_*

**Algorithm A3: Feature extraction and model training**

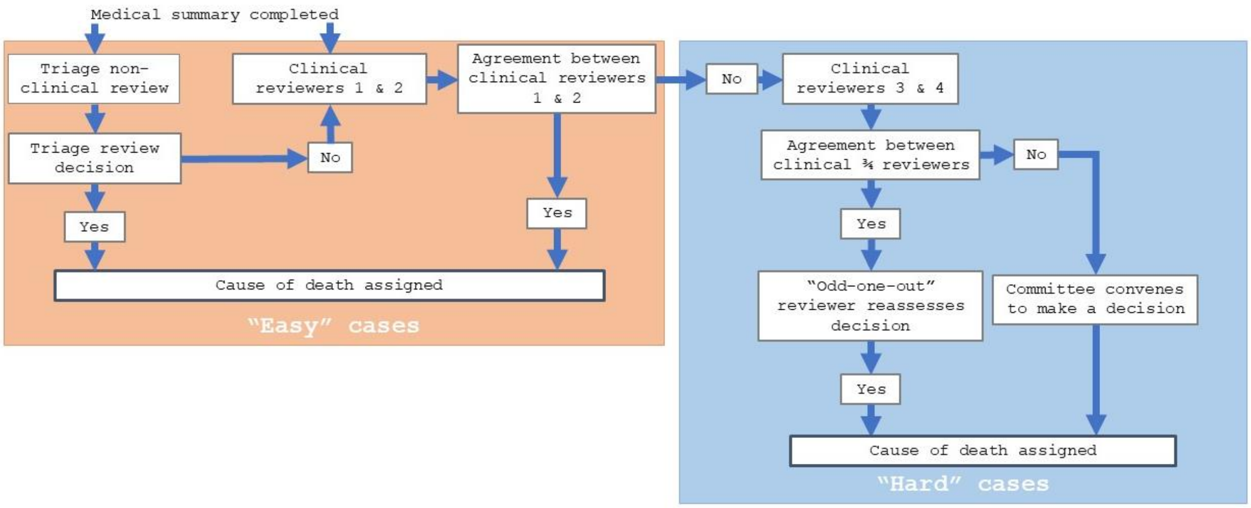

### ESM_2: Interpretable Vignette – Non prostate cancer death

#### Interpretable Vignette Classification for Cause of Death Review

Study ID number : [redacted]
Date of death [redacted]
Date of diagnosis : [redacted]

Predicted death code: 2 (0.71)
Actual death code: 2
COD route: 5

Feature contribution legend

bone scan — Largest negative contribution
uptake right lesion — Smallest contributions
hormone — Largest positive contribution

**Clinical features at diagnosis**

Gleason Score at diagnosis (with dates): [redacted]/2010 ? Gleason 3+5=8

Clinical stage (TNM): [redacted]/2009 ? ?Benign?, TRUS volume = 60cc

Pathological stage (TNM): -

Co-morbidities with dates of diagnosis: Unknown diagnosis dates: Hypertension and high cholesterol Abdominal aortic aneurysm ? under surveillance

Other primary cancers with dates of diagnosis: -

PSA level at diagnosis with dates: [redacted]/2010 - 119ug/L

Radiological evidence of local spread at diagnosis: [redacted]/2010 ? MRI Pelvis Prostate: There is definite prostate hypertrophy noted. No definite extra-prostatic disease seen. The peripheral zones are difficult to visualise. Seminal vesicles are normal. No significant pelvic lymphadenopathy seen. No evidence of bone metastases seen in the lumbar spine. Conclusion: Radiological staging would be T2, N0, M0.

Radiological evidence of metastases at diagnosis: [redacted]/2009 ? No evidence of bony metastases

**Clinical features at diagnosis**

Initial treatments (dates): Radical Radiotherapy 01 [redacted]? [redacted]2010 plus 2 years of Hormone therapy: CPA started [redacted]2010 for 3 weeks, Prostap started [redacted]/10 ? completed [redacted]/2012

Hormone therapy (start date): -

Maximum androgen blockade (start date): -

Orchidectomy (date): -

Chemotherapy (start date): -

Treatment for complications of treating prostate cancer with dates (if available): -

**Clinical features at diagnosis**

Serial PSA levels (dates): [redacted]/2011 ? 0.3ug/L

[redacted]/2012 - <0.2ug/L

[redacted]/2012 - <0.2ug/L

---

Serum testosterone: -

---

Radiological evidence of metastases: See scans below.

---

Other indications or complications of disease progression: [redacted]/2010 ? DRE Small feeling prostate, soft, symmetrical nodes.

---

Date of recurrence following radical surgery or radiotherapy: -

---

Palliative care referrals and treatments:

---

**Clinical features at diagnosis**

---

Treatment/ admission for co-morbidity with dates (if available): [redacted]/2012 CT Thorax: There are multiple solid masses in the liver in keeping with metastases. There is a circumferential tumour at the distal body of the stomach with infiltration into the surrounding fat and probable involvement of the adjacent gastro epiploic vessels. No enlarged nodes. Conclusion: Gastric cancer T4 N0 M1

[redacted]/2014 ? CT Thorax, abdomen and pelvis: Disease progression with increase in the primary gastric mass as well as in the liver metastases.

[redacted]/2015 ? CT Angiogram pulmonary: No PE. There are new deposits in the lungs and ascites in keeping with progressive disease.

---

**Clinical features at diagnosis**

---

Symptoms in last 3-6 months (i.e. bone pain, weight loss, cachexia, loss of appetite, obstructive uraemia): See below

---

Last consultation: speciality & date: [redacted]2015 ? Medical Oncology

---

Was a DS1500 report issued?: -

---

Post mortem findings: Not seen

### ESM_3: Interpretable Vignette –Prostate cancer death

**Interpretable Vignette Classification for Cause of Death Review**

Study ID number : [redacted]
Predicted death code: 1 (0.16)

Date of death : [redacted]
Actual death code: 1

Date of diagnosis : [redacted]
COD route: 1

Feature contribution legend

docetaxel — Largest negative contribution

smaller contains — Smallest contributions

hormone — Largest positive contribution

**Clinical features at diagnosis**

Gleason Score at diagnosis (with dates): [redacted]/2004 Gleason 4+5 =9 in 4/4 cores. Perineural invasion. No extraprostatic invasion.

Clinical stage (TNM): [redacted]/2004 DRE ? T3 disease

Pathological stage (TNM): -

Co-morbidities with dates of diagnosis: None

Other primary cancers with dates of diagnosis: None

PSA level at diagnosis with dates: [redacted]2004 110 ug/L

Radiological evidence of local spread at diagnosis: [redacted]2004 TRUS ? malignant looking gland.

[redacted]/2004 Flexible cystoscopy ? no evidence of intravesical abnormality, but prostate was friable and bled on touch at passing the cystoscope

Radiological evidence of metastases at diagnosis: [redacted]2004 Bone scan ? raised uptake at right scapula, ribs, L4, several areas in the pelvis (both sides), possible area left upper femur. Appearances consistent with bone metastases.

**Clinical features at diagnosis**

Initial treatments (dates): [redacted]2004 Hormones

[redacted]/2005 Starts steroids

[redacted]/2007 Starts bisphosphonates (4 cycles Zoledronate)

[redacted]2007 Starts chemotherapy (4 cycles Taxotere)

[redacted]/2007 Starts Diethylstilboestra

Hormone therapy (start date): [redacted]2004 Cyproterone Acetate

[redacted]/2004 Zoladex

Maximum androgen blockade (start date): -

Orchidectomy (date): -

Chemotherapy (start date): [redacted]/2007 First cycle Taxotere chemotherapy (4 in total)

Treatment for complications of treating prostate cancer with dates (if available): 4 inpatient admissions following chemotherapy treatments

[redacted]/2007 - [redacted]/2007: severe diarrhoea and dehydration.

[redacted]/2007 - [redacted]/2007: diarrhoea, nausea, vomiting

[redacted]/2007 - [redacted]/2007: general weakness, lethargy

[redacted]/2007 - [redacted]/2007: nausea and vomiting

**Clinical features at diagnosis**

Serial PSA levels (dates): [redacted]/2005 0.9 ug/L

[redacted]/2005 70.2 ug/L

[redacted]/2005 223 ug/L

[redacted]/2006 14.8 ug/L

[redacted]/2006 240 ug/L

[redacted]/2007 235 ug/L

[redacted]/2007 247 ug/L

[redacted]/2007 326 ug/L

[redacted]/2007 440 ug/L

[redacted]/2007 622 ug/L

[redacted]/2007 449 ug/L

[redacted]/2007 440 ug/L

[redacted]/2007 398 ug/L

Serum testosterone: -

Radiological evidence of metastases: [redacted]/2007 Bone scan ? Widespread focal and diffuse areas of raised uptake in the axial skeleton and right proximal femur. Marked increase in extent of uptake since the previous scan ([redacted]-04). Extensive bone metastases, progressive disease.

[redacted]/2007 Bone scan ? multiple focal and diffuse areas of tracer uptake in the ribs, thoracic and lumbar spine, pelvis, skull and right pxoximal femur. Appearances are largely unchanged since the previous scan ([redacted]-07).

Other indications or complications of disease progression: [redacted]/2005 Diagnosis - hormone resistant prostate cancer. Symptomatic. Starts steroids

[redacted]/2007 Disease progression - 4 months of chemotherapy PSA responding, but general condition deteriorating (loosing weight

| |
|---|
| and appetite). Stops Taxotere to improve QOL. Starts Diethylstilboestral. |
| Date of recurrence following radical surgery or radiotherapy: - |
| Palliative care referrals and treatments: |
| **Clinical features at diagnosis** |
| Treatment/ admission for co-morbidity with dates (if available): N/a |

| **Clinical features at diagnosis** |
|---|
| Symptoms in last 3-6 months (i.e. bone pain, weight loss, cachexia, loss of appetite, obstructive uraemia): Nausea, vomiting, bone pain, tiredness |
| Last consultation: speciality & date: [redacted]/2007 - [redacted]/2008 Inpatient (Hospice) |
| Was a DS1500 report issued?: - |
| Post mortem findings: - |
